# Learning From the Past: Medical School Experiences Predict Meaning in Work and Imposter Phenomenon Among Residents

**DOI:** 10.1101/2025.02.11.25322077

**Authors:** Kelsey M. Carpenter, Christopher M. Falco, Sabrina Menezes, Jessica B. B. Diaz, Jessica C. Wells, Gregory Guldner, Jason T. Siegel

## Abstract

**Introduction:** Endeavors to bolster resident well-being may benefit from a greater understanding of how associated psychological states are shaped across various stages of medical education. To this end, the current study aimed to determine if perceived psychological safety and exposure to negative aspects of the hidden curriculum during medical school predicted meaning in work and imposter phenomenon among medical residents transitioning into their new programs.

**Methods:** All incoming residents at a multi-institutional hospital organization were administered a survey link via email as part of a larger set of studies. Participants in this specific study (*n* = 237) completed measures assessing their perceptions of both positive and negative medical school experiences (i.e., psychological safety and hidden curriculum exposure) as well as their current feelings about themselves and their work (i.e., meaning in work and imposter phenomenon). Data collection took place during the summer of 2022.

**Results:** Perceptions of psychological safety during the last two years of medical school were positively related to meaning in work and negatively related to imposter phenomenon among incoming residents. Perceptions of exposure to negative aspects of the hidden curriculum predicted imposter phenomenon, but no link was observed between hidden curriculum exposure and meaning in work.

**Discussion:** Feelings related to previous learning environments (i.e., medical school) may still be relevant during the transition into residency. Next steps include exploring potential causal linkages between these variables as well as expanding the scope and operationalization of psychological states and experiences considered across time points during the medical training journey.

## Introduction

Medical residents undergo an arduous training process that presents numerous threats to their well-being.^1^ Despite growing efforts to curb distress within this population,^2^ further action is needed.^3^ Research within this realm often focuses on the residency stage in isolation. However, residents do not enter their programs as blank slates; rather, their well-being is likely impacted by their current environment^4^ as well as prior experiences in their educational journey.^5,6^ Successfully addressing resident well-being could require greater consideration of how residents’ current thoughts and feelings may have roots in their previous experiences, namely, those in medical school. Residents transitioning from medical school environments that they perceive as more supportive may be better equipped to maintain well-being during residency, whereas those from less affirming environments could face challenges before even setting foot into a residency program. To this end, the current research incorporates consideration of medical school into an investigation of the psychological states of incoming residents, determining how perceived experiences during medical school may predict aspects of their well-being during this vital transitional stage.

### Medical School Experiences

To explore potential ties between medical school experiences and incoming resident well-being, we examined two variables tied to the clinical learning environment: psychological safety and exposure to the hidden curriculum. Psychological safety refers to “a shared belief that the team is safe for interpersonal risk taking,”^7(p354)^ including disclosing mistakes, raising concerns, or asking for help, without facing consequences such as embarrassment or punishment.^8^ Within the healthcare context, the presence of psychological safety has been linked to favorable perceptions of the clinical learning environment^9^ and deeper immersion in the learning process.^10^ Residents who reflect on their time in medical school and perceive that their clinical rotations took place in psychologically unsafe learning environments may harbor residual feelings and expectations for residency distinct from those held by others who felt more secure during medical school. The hidden curriculum generally refers to influences on learning beyond the lessons that are intentionally passed on through formal channels.^11–13^ This construct encompasses a broad set of forces shaping educational experiences, including those ingrained within an organization’s culture as well as the transmission of norms, expectations, and values through interpersonal interactions and modeling of behavior.^14,15^ Considering some of the more detrimental manifestations of the hidden curriculum, medical students may pick up on underlying rules about when it is acceptable or not to speak up,^16^ observe questionable behavior among residents and attendings, such as the disparagement of patients behind their backs,^16^ or encounter situations that challenge their idealistic views of medicine.^17^ Exposure to these negative dimensions of the hidden curriculum could hold lasting consequences for learners regarding their perceptions of their work, confidence in their ability to become competent medical professionals, and overall well-being.

### Psychological States of Incoming Residents

Efforts to maximize well-being during residency may be the most lucrative if training programs are designed to promote flourishing from the start, rather than intervening as issues arise down the line.^18^ As such, it is imperative to understand the mindsets of the residents as they transition into their new roles, determining their support needs at this stage before they are fully immersed into their new environments. Although numerous factors are often considered when investigating well-being, we focused on one positive and one negative construct especially pertinent to this stage: meaning in work and imposter phenomenon. Meaning in work is a global perception that individuals have about their work, which may encompass beliefs about the societal contributions of their vocation^19^ as well as their work’s influence on personal growth^20^ and level of congruence with their personal values.^21^ This construct is especially relevant for resident physicians as they start to develop their identities as practicing medical professionals, tasked with alleviating pain and disease while encountering numerous structural factors that may detract from their ability to see the positive impact of their day-to-day tasks.^22^ Given the accumulating evidence for associations between greater meaning in work and lower levels of depression^20^ and increased specialty and career satisfaction^23–25^ as well as consideration of its ties to burnout,^26^ maintaining a sense of meaning in work is likely a key contributor to resident well-being. Imposter phenomenon refers to “an internal experience of intellectual phoniness.”^27(p. 241)^ In line with other work, we intentionally use the term “phenomenon” rather than “syndrome” to reinforce the notion that this experience is often generated from or exacerbated by environmental factors rather than an issue that arises unprompted within the individual.^4,28^ Imposter phenomenon has been documented across the spectrum of medical training, from medical residents in various specialties^29,30^ to practicing physicians.^31^ It has also been linked to impaired well-being, hindered job performance,^32^ and burnout.^31^ Given the often competitive undertone of medical learning environments,^33^ the public nature of learning,^34^ and pressure to stay up-to-date with a rapidly expanding body of knowledge,^35^ new residents may be especially prone to imposter phenomenon. Accordingly, our study aimed to determine how residual experiences from medical school (i.e., psychological safety and exposure to negative aspects of the hidden curriculum) relate to meaning in work and imposter phenomenon during the transition into residency.

### Current Study

We surveyed incoming residents at a nationwide hospital corporation with facilities that sponsor over 5,000 residents to better understand how medical residents may be affected by their experiences in medical school. We hypothesized that (1) lower psychological safety and (2) greater exposure to negative aspects of the hidden curriculum in medical school would be related to (a) lower meaning in work and (b) higher imposter phenomenon when entering residency.

## Methods

We conducted this research as part of a larger data collection effort targeting incoming medical residents across a multi-institutional hospital network. The online survey was administered in the summer of 2022 via a link emailed to incoming residents by administrators at the hospital organization. The hospital network’s Institutional Review Board deemed this study exempt from formal review.

### Participants

The survey was sent out to 1,674 incoming residents. In total, 1,339 individuals completed at least 75% of their assigned survey, indicating a response rate of 79.9%. The data collection involved four independent studies, and 297 participants were sorted into the survey path used in the current study. After listwise deletion of responses due to failure to provide informed consent, missing data, and removal of outliers whose Mahalanobis distance exceeded the 99^th^ percentile, the final sample size was 237.

### Materials

As part of the survey, incoming medical residents completed four self-report measures: two indicating their perceptions of their medical school experiences, and two indicating their current feelings as they started the transition into residency. Respondents also provided demographic information.

#### Psychological safety

Psychological safety was measured using Edmondson’s^7^ seven-item Likert scale (strongly disagree/strongly agree). The items were adapted to further tailor the scale to this research context, instructing respondents to reflect back on their experiences during their clinical rotation teams. For example, the item “Members of this team are able to bring up problems and tough issues” was altered to “Members of clinical rotation teams were able to bring up problems and tough issues.” In line with Kerrissey and colleagues,^36^ we examined psychological safety at the individual level rather than at the group level as our research aim was to consider the influence of the residents’ personal perceptions of psychological safety rather than to indicate the shared beliefs held by all members of their clinical rotation teams, though these may overlap. The response set was also expanded from five to seven points to capture more variance.

#### Hidden curriculum

The current research used 26 items adapted from Billings and colleagues’^37^ hidden curriculum scale to assess perceptions of exposure to the hidden curriculum in medical school. The items, which were condensed due to survey spatial constraints, focused on negative manifestations of the hidden curriculum, prompting incoming residents to indicate the frequency with which they felt humiliated, witnessed unprofessional conduct, or observed medical record falsification among various groups during their clinical rotations (e.g., “Please answer with how often you typically witnessed the following during your clinical experiences in your last two years of medical school… Felt humiliated by Faculty/Medical staff”). Respondents answered on a five-point scale (never/more than six times).

#### Meaning in work

The Work and Meaning Inventory (WAMI)^20^ assessed meaning in work among the incoming residents. The WAMI is a 10-item, five-point Likert scale (absolutely untrue/absolutely true) with three subscales: positive meaning (e.g., “I have found a meaningful career”), meaning-making through work (e.g., “My work helps me make sense of the world around me”), and greater good motivations (e.g., “The work I do serves a greater purpose”).

#### Imposter phenomenon

The Clance Imposter Phenomenon Scale (CIPS)^38^ assessed the residents’ current levels of imposter phenomenon. This five-point Likert scale (not at all true/very true) consists of 20 items (e.g., “Sometimes I feel or believe that my success in life or in my job has been the result of some kind of error”), with higher scores indicating more elevated feelings of imposterism.

## Results

All data met the assumption of homoscedasticity, χ^2^(1) = 0.09, *p =* .771, as well as the assumption of linearity. Although a Henze Zirkler test indicated the data were not multivariate normal, HZ = 2.33, *p* < .001, the qq-plot confirmed the violation was minor. Considering regression is robust to minor violations of non-normality, we proceeded with our planned analysis (see Table 1 for descriptive data).

**Table 1.** Descriptive Statistics.

|  | Psychological Safety | Hidden Curriculum | Meaning in Work | Imposter Phenomenon |
| --- | --- | --- | --- | --- |
| N | 237 | 237 | 237 | 237 |
| Missing | 0 | 0 | 0 | 0 |
| Mean | 5.12 | 16.66 | 43.62 | 50.02 |
| Median | 5.14 | 14.00 | 45.00 | 48.00 |
| Standard Deviation | 0.91 | 15.94 | 5.75 | 16.66 |
| Minimum | 2.71 | 0.00 | 26.00 | 20.00 |
| Maximum | 7.00 | 71.00 | 50.00 | 100.00 |
| Cronbach's Alpha | 0.73 | 0.95 | 0.93 | 0.95 |

T-tests comparing white residents and residents of color found no significant differences in meaning in work, *t*(231) = 0.34, *p* = .737 or imposter phenomenon, *t*(231) = 0.67, *p* = .503. There were also no significant gender differences in meaning in work, *F*(3,230) = 2.37, *p* = .071 or imposter phenomenon, *F*(3,230) = 1.18, *p* = .317. As such, we did not control for either demographic category in our final specified models.

Model 1 tested the relationship between psychological safety and meaning in work. This relationship was significant, β = .27, *p* < .001, 95% CI [.15, .39]; psychological safety had a significant positive relationship with meaning in work, fully supporting Hypothesis 1a. Model 2 returned a significant negative relationship between psychological safety and imposter phenomenon, β = -.31, *p* < .001, 95% CI [-.43, -.18]. As such, Hypothesis 1b was also fully supported (see Table 2 for all regression results).

**Table 2.** Simple Regression Results.

| Outcome | Predictor | Estimate | SE | 95% CI | | t | p | $\beta$ | R<sup>2</sup> |
| --- | --- | --- | --- | --- | --- | --- | --- | --- | --- |
|  |  |  |  | Lower | Upper |  |  |  |  |
| Meaning in Work | Intercept | 34.83 | 2.07 | 30.76 | 38.91 | 16.84 | <.001 |  | 0.07 |
|  | ps | 1.72 | 0.40 | 0.93 | 2.50 | 4.32 | <.001 | 0.27 |  |
| Meaning in Work | Intercept | 44.14 | 0.54 | 43.07 | 45.20 | 81.79 | <.001 |  | 0.01 |
|  | hc | -0.03 | 0.02 | -0.08 | 0.02 | -1.31 | 0.19 | -0.09 |  |
| Imposter Phenomenon | Intercept | 78.82 | 5.93 | 67.13 | 90.50 | 13.29 | <.001 |  | 0.09 |
|  | ps | -5.62 | 1.14 | -7.87 | -3.38 | -4.93 | <.001 | -0.31 |  |
| Imposter Phenomenon | Intercept | 45.43 | 1.51 | 42.44 | 48.41 | 29.99 | <.001 |  | 0.07 |
|  | hc | 0.28 | 0.07 | 0.15 | 0.41 | 4.19 | <.001 | 0.26 |  |

*Note.* ps = psychological safety; hc = hidden curriculum

Models 3 and 4 tested the effects of hidden curriculum on meaning in work and imposter phenomenon. Hypothesis 2a was not supported, as there was no relationship between hidden curriculum and meaning in work, β = -.09, *p* = .190, 95% CI [-.21, .04]. However, Hypothesis 2b was fully supported, as there was a significant positive relationship between hidden curriculum and imposter phenomenon, β = .26, *p* < .001, 95% CI [.14, .39]. Please see Figure 1 for a visualization of all model results.

**Figure 1.**
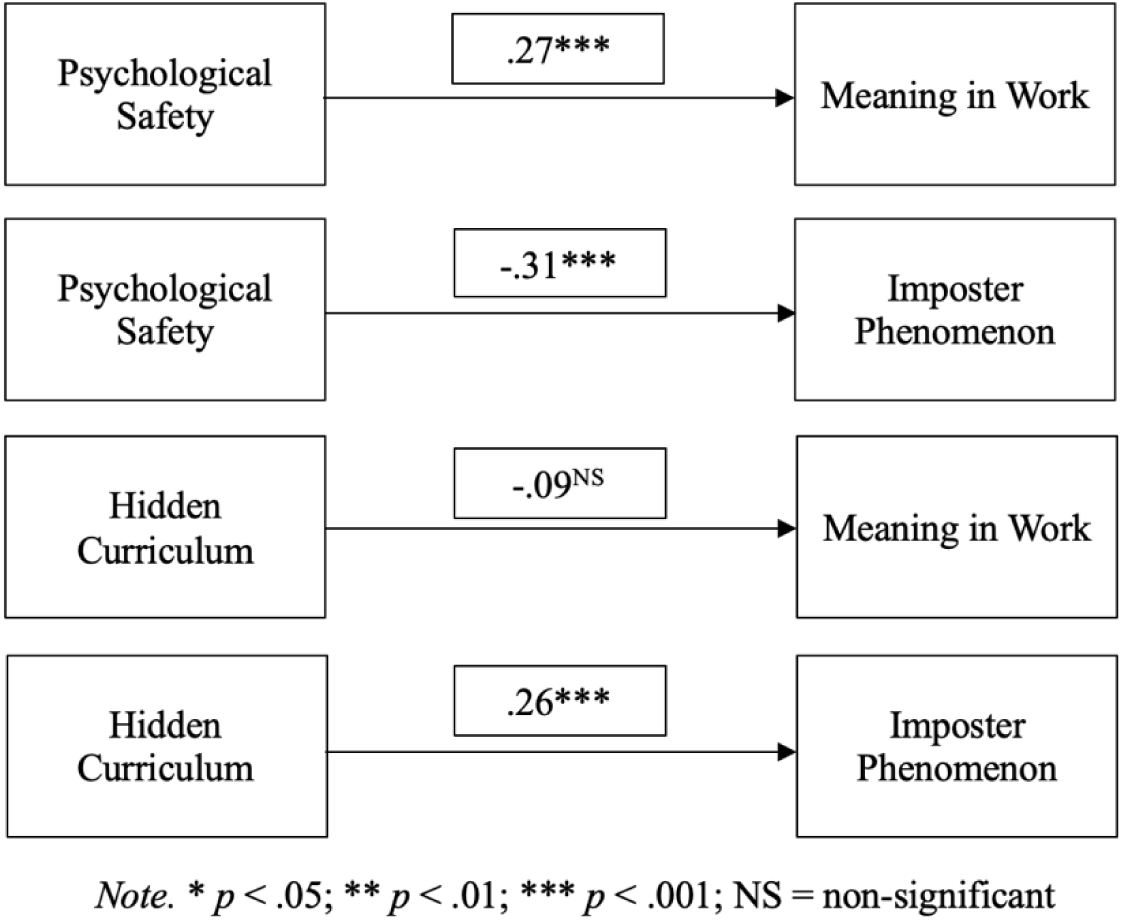
Visualization of Model Results.

## Discussion

Complementing widespread efforts to foster well-being among medical residents and mitigate associated barriers, we surveyed incoming residents at a large hospital network to better understand how their medical school experiences may play a role in their current states. We asked transitioning residents to reflect upon their final years of medical school and report their perceptions of psychological safety and exposure to the hidden curriculum, anticipating that these factors would predict their current feelings regarding meaning in work and imposter phenomenon. Most of our initial hypotheses were supported, representing a rare demonstration of how residual feelings about previous learning environments (i.e., medical school) may still be relevant during the transition into residency.

In line with our hypotheses, perceptions of psychological safety during the last two years of medical school predicted meaning in work among the incoming medical residents. This aligns with prior qualitative literature indicating a relationship between safe learning environments and a sense of belonging, such as McClintock and colleagues’^18^ examination among fourth-year medical students and Hipp and colleagues’^39^ determination of the importance of a collegial work environment in enhancing meaning in work among residents and fellows. Psychological safety was also supported as a predictor of imposter phenomenon, such that when residents reported having felt more psychologically safe in clinical rotation teams, their current feelings of imposterism were lower. One proposed mechanism that may explain this link is engagement in learning.^40^ For instance, Bynum and colleagues^41^ suggest that because medical training inherently requires risk, the presence or absence of feelings of psychological safety may determine whether learners immerse themselves further in the learning process or become detached. Although exploring engagement in the learning process was beyond the scope of the current study, this could be an important avenue for future research.

Contrary to our predictions, a relationship between hidden curriculum and meaning in work was not observed. We were surprised by these findings, especially given previous literature suggesting that certain components of the hidden curriculum, such as witnessing the mocking of patients,^16^ may contribute to discordance between individuals’ imagined experiences as medical professionals and their day-to-day reality.^17,42^ However, as noted by Higashi et al.,^43^ students may not passively absorb all messages transmitted through the hidden curriculum; rather, as critical thinkers, they may reject specific components that do not align with their personal values and goals. Exposure to negative aspects of the hidden curriculum did, however, serve as a predictor of imposter phenomenon among the incoming medical residents. If the presence of a negative hidden curriculum contributes to inhibitions in learning behavior, such as learning that speaking is not always acceptable,^16,44^ it stands to reason that medical students may hold back questions, resulting in perceived knowledge gaps and contributing to fears of incompetence. As such, increased focus on uncovering the negative messages transmitted through the hidden curriculum and amplifying the positive ones is crucial.^15^

This research has limitations, many of which can be overcome with further inquiries into the topics examined within this study. For instance, the operationalization of our chosen variables may fail to paint a complete picture of the relationships between the constructs. As an example, our investigation explored specific negative aspects of the hidden curriculum, focusing on experiences such as humiliation and witnessing unprofessional conduct. Ideally, future work will expand this research by further considering the positive aspects that may comprise the hidden curriculum, such as the modeling of professionalism, empathy, and self-care.^45^ Another noteworthy aspect of our investigation pertains to the specific career stage assessed during this study: the transition between medical school and residency. Although this time period is essential to explore, it is possible that negative feelings associated with this role adjustment (e.g., anxiety) could contribute to heightened negative feelings as respondents reflect back on their previous states or report their current beliefs. It also may be the case that feelings related to meaning in work and imposter phenomenon shift as the residents become more immersed in their new positions. Ideally, future studies can delve more into the impact of temporality, including surveying medical students before graduation and following up with the residents after they have had more time to acclimatize to their environments.

Overall, these findings indicate that perceived experiences during medical school are tied to the psychological states of incoming residents. Beyond providing insight into critical emotional experiences during this transitional period, research that considers related medical school factors could be imperative for understanding how to support the well-being of residents most effectively, accounting for both current issues as well as potential roots in the past. It is possible that interventions targeting imposter phenomenon, for instance, may be strengthened if they also consider how previous environments that felt psychologically safe or unsafe may have contributed to emotions such as shame or unrealistic beliefs about the required abilities to be a competent physician. Moreover, if surveys such as the one conducted for the current study are implemented, program directors could use information about medical school experiences, which may be less susceptible to social desirability bias, to predict which residents might have the greatest need for intervention. Further research exploring these variables in more depth, including studies with longitudinal designs, different operationalizations of these variables, or incorporation of other factors, may allow for developing interventions or establishing environments that promote flourishing while minimizing harm.

By examining perceptions of medical school experiences and the current psychological states of physicians transitioning into their resident roles, our findings reinforce the notion that attempts to address resident well-being may benefit from considering broader contextual influences such as previous learning environments. Specifically, these results contribute to a more robust understanding of the lasting impact of medical school on incoming residents and how their past may predict key emotional states during this transitional period. To be sure, no amount of individual intervention in medical school or residency is likely to cancel out the detrimental impact of harmful learning environments.^41^ Cultural and systemic change must be prioritized,^46^ including addressing structural factors contributing to distress as well as working to establish healthier norms that still align with the values and identities of the physicians.^4^ In the meantime, however, it is essential to learn how to promote flourishing at all stages of medical education despite the given constraints to well-being.^5,46^ Collective commitment to identifying and addressing modifiable stressors in the training process,^47^ establishing psychologically safe learning environments,^48^ and the provision of concrete support based upon the voiced needs of the residents may help reduce the obligation to sacrifice their own well-being in pursuit of aiding others.

## Data Availability

All data produced in the present study are available upon reasonable request to the authors

## Structured Disclosures

### Acknowledgements

The authors wish to thank Bruce Deighton for his support.

### Ethical approval

This study was deemed exempt by the institutional review board at the hospital network (6/15/2022, approval number 2022-496).

### Disclaimers

This research was supported (in whole or in part) by HCA Healthcare and/or an HCA Healthcare affiliated entity. The views expressed in this publication represent those of the authors(s) and do not necessarily represent the official views of HCA Healthcare or any of its affiliated entities.

### Previous presentations

These findings have been distributed as an internal report at the hospital network.

### Data

The authors have permission from Claremont Graduate University and HCA Healthcare to distribute these data.

